# Rotational Thromboelastometry predicts care level in COVID-19

**DOI:** 10.1101/2020.06.11.20128710

**Authors:** Lou M. Almskog, Agneta Wikman, Jonas Svensson, Michael Wanecek, Matteo Bottai, Jan van der Linden, Anna Ågren

## Abstract

**Background:** High prevalence of thrombotic events in severely ill COVID-19 patients have been reported. Pulmonary embolism as well as microembolization of vital organs may in these individuals be direct causes of death. The identification of patients at high risk of developing thrombosis may lead to targeted, more effective prophylactic treatment.

**Objectives:** To test whether Rotational Thromboelastometry (ROTEM) indicates hypercoagulopathy in COVID-19 patients, and whether patients with severe disease have a more pronounced hypercoagulopathy compared with less severely ill patients.

**Methods:** The study was designed as a prospective observational study where COVID-19 patients over 18 years admitted to hospital were eligible for inclusion. Patients were divided into two groups depending on care level: 1) regular wards or 2) wards with specialized ventilation support. ROTEM was taken after admission and the data were compared with ROTEM in healthy controls.

**Results:** The ROTEM variables Maximum Clot Firmness (EXTEM-/FIBTEM-MCF) were higher in COVID-19 patients compared with healthy controls (p<0.001) and higher in severely ill patients compared with patients at regular wards (p<0.05). Coagulation Time (EXTEM-CT) was longer and Clot Formation Time (EXTEM-CFT) shorter in COVID-19 patients compared with healthy controls. Our results suggest that hypercoagulopathy is present in hospitalized patients with mild to severe COVID-19 pneumonia.

**Conclusion:** ROTEM variables were significantly different in COVID-19 patients early after admission compared with healthy controls. This pattern was more pronounced in patients with increased disease severity, suggesting that ROTEM-analysis may be useful to predict thromboembolic complications in these patients.

## Background

Corona virus disease 2019 (COVID-19), caused by the novel corona virus SARS-CoV-2, has the last few months taken hold of the world and spread as a global pandemic with presently more than 8.0 million cases ^1^. The lungs are the main target organ for COVID-19, and patients who become critically ill generally suffer from respiratory distress leading to difficulties in ventilation and oxygenation. However, the respiratory symptoms in COVID-19 display atypical features, which have challenged the conventional mechanical ventilation strategies ^2^.

A recent Dutch study of 184 COVID-19 positive patients treated at intensive care units (ICUs) showed a cumulative incidence of thrombotic events of 49%, mainly pulmonary embolism ^2^. Autopsy reports have confirmed and even exceeded this high rate of thrombosis ^3,4^, and have also shown a high prevalence of microthrombosis in the small veins of the lungs ^5^. Thromboembolic complications may be a direct cause of death in COVID-19 ^3,4^ and in addition, the loss of perfusion caused by thrombi in the lungs will impair pulmonary gas exchange independently of the direct tissue damage caused by the viral pneumonia. This will contribute to the respiratory failure that is the main cause of critical disease in patients with COVID-19.

Elevated levels of fibrin degradation products (e.g. D-dimer) have consistently been reported as a strong prognostic factor associated with poor outcome in patients with COVID-19 ^6,7^. This suggests a general coagulation activation and secondary hyperfibrinolysis. However, the D-dimer increase is not evident in early stages of the disease ^7^, limiting its usefulness as a prognostic marker.

Maximum clot firmness (MCF) may be calculated using Rotational Thromboelastometry (ROTEM) and is considered a good marker for hypercoagulopathy ^8^. Theoretically, ROTEM variables may be affected earlier during the disease course compared with D-dimer and may be of greater value as a predictor of disease severity. Studies assessing ROTEM in COVID-19 patients treated at ICUs have shown elevated MCF values ^9,10^. However, no ROTEM data at earlier stages of the disease have, to our knowledge, yet been reported.

The aim of this study was to assess the presence of coagulopathy, measured by ROTEM, in COVID-19 patients at hospital admission.

## Materials and methods

### Study design

The study was designed as a prospective observational study and was approved by the Swedish Ethical Review Authority (D-nr 2020-01875). Patients over 18 years of age, who tested positive for COVID-19 and were considered in need of hospital care were eligible for inclusion in the study. Recruitment of patients commenced May 8th, 2020, and is planned to continue for three months in total. Here we report on subjects included up to May 31st, 2020.

All patients were included at Capio St Göran’s Hospital (StG) in Stockholm, Sweden. Patients presenting at StG Emergency Room with confirmed SARS-CoV-2 infections and in need of hospitalization are admitted either to regular wards with possibility of low-flow oxygen therapy (henceforth “Regular ward”) or to wards with possibilities of specialized ventilation support; either non-invasive ventilation in intermediate wards (Biphasic Positive Airway Pressure (BiPAP) or High-Flow Oxygen Therapy (HFOT)) or to the Intensive Care Unit where, in addition to BiPAP and HFOT, mechanical ventilation support is provided (henceforth “Specialized wards”).

Conventional coagulation tests (CCTs): D-dimer, P-fibrinogen, Activated Partial Thromboplastin Time (APTT), International Normalized Ratio (INR), Antithrombin and Platelet count as well as ROTEM were drawn from all included patients as soon after admission to hospital as possible (median within one day after admission). ROTEM test results were blinded for clinicians, but CCTs were not. Aside from these blood tests, nothing was changed in the routine care of included patients.

Anticoagulant treatment after admission were categorized according to 1) low prophylactic dose LMWH = 75 IE/kg/24h; 2) high prophylactic dose LMWH = 150 IE/kg/24h; 3) treatment dose LMWH ≥ 200 IE/kg/24h; or 4) pre-existing anticoagulant medication, e.g. new oral anticoagulants.

A ROTEM sigma (Tem Innovations GmbH, Germany) was used for the thromboelastometric analyses. ROTEM sigma is a fully automated system with proven ROTEM technology, where pipetting and test preparation are not required. Analyzing time is 45-60 min for each test. All blood samples were analyzed at StG laboratory within four hours after they were drawn.

ROTEM data collected from healthy individuals 2011-2012 for a previously published study were used as controls. The original publication contains further details ^11^.

### ROTEM-tests

Two of four ROTEM-variables are presented here: i) extrinsically activated assays with tissue factor (EXTEM); ii) with tissue factor and platelet inhibitor cytochalasin D (FIBTEM)^12^. EXTEM tests the extrinsic coagulation pathway. FIBTEM blocks the platelet contribution to clot formation, leaving only the impact of fibrin formation and polymerization. Coagulation Time (CT) is the time (in seconds) from test start until an amplitude of 2 mm is reached, giving information about coagulation activation / initiation. Clot Formation Time (CFT) is the time (in seconds) between 2 mm amplitude and 20 mm amplitude, giving information about clot propagation. Maximum Clot Firmness (MCF) is the maximum amplitude (in millimeters) reached during the test, giving information about clot stability. A short EXTEM-CFT and an increased EXTEM-MCF and/or FIBTEM-MCF suggest a hypercoagulable state.

### Statistical analysis

All continuous variables are presented as median and interquartile range (IQR). Kruskal-Wallis tests were used to test whether ROTEM data for COVID-19 patients differed depending on care level (Regular ward; Specialized wards), or from healthy controls. If the tests were significant, pairwise two-sided Wilcoxon tests were performed. Spearman’s correlation coefficients were calculated to assess associations between variables. P-values below 0.05 were considered statistically significant. R version 4.0.0 was used to perform statistical analysis and visualizations.

## Results

Sixty COVID-19 positive patients and 89 healthy controls were included in the analysis. *Table 1* presents patient characteristics. Comorbidities were common within the COVID-19 patient group, where 42% of patients had a prior diagnosis of hypertension and 28% of diabetes. 48 patients (80%) received prophylactic anticoagulant treatment (ACTr) after admission, and in 36 (60%) patients this treatment was administered before ROTEM was analyzed. Of patients with ACTr before ROTEM, 19 received low dose prophylaxis with Low Molecular Weight Heparin (LMWH), 7 had high dose prophylaxis with LMWH, 5 had treatment dose LMWH and 5 continued their regular anticoagulant medication.

**TABLE 1.** Baseline characteristics of COVID-19 patients and healthy controls.

|  | COVID-19 |  |  | Healthy controls |
| --- | --- | --- | --- | --- |
|  | Regular ward | Specialized wards | Total |  |
|  | N=40 | N=20 | N=60 | N=89 |
| Gender male, N (%) | 28 (70%) | 12 (60%) | 40 (67%) | 49 (55%) |
| Age, years, median (IQR) | 61 (51-74) | 62 (55-66) | 61 (52-71) | 49 (37 - 56) |
| BMI, kg/m<sup>2</sup>, median (IQR) | 26 (24-32) | 28 (25-32) | 27 (24-32) |  |
| Previous thromboembolic disease, N(%) | 4 (10%) | 2 (10%) | 6 (10%) |  |
| Anticoagulant treatment at inclusion, N (%) | 12 (30%) | 4 (20%) | 16 (27%) |  |
| Diabetes, N (%) | 8 (20%) | 9 (45%) | 17 (28%) |  |
| Smoking, N (%) | 2 (5%) | 3 (15%) | 5 (8%) |  |
| COPD/asthma, N (%) | 6 (15%) | 4 (20%) | 10 (17%) |  |
| Hypertension, N (%) | 13 (33%) | 12 (60%) | 25 (42%) |  |
| Cardiovascular disease, N (%) | 5 (13%) | 4 (20%) | 9 (15%) |  |
| Malignancy, N (%) | 4 (10%) | 4 (20%) | 8 (13%) |  |
| Other diseases, N (%) | 9 (23%) | 6 (30%) | 15 (25%) |  |
| Days with COVID symptoms at inclusion, median (IQR) | 10 (7-14) | 12 (9-16) | 11 (7-14) |  |
| Days at hospital at inclusion, median (IQR) | 1 (1-2) | 2.5 (1-5) | 1 (1-2) |  |
| Thrombosis at inclusion, N (%) | 2 (5%) | 4 (20%) | 6 (10%) |  |
| Anticoagulant prophylaxis before ROTEM analysis, N (%) | 21 (53%) | 15 (75%) | 36 (60%) |  |

IQR=Interquartile Range. BMI=Body Mass Index. COPD=Chronic Obstructive Pulmonary Disease.

*Table 2* presents laboratory test results in healthy controls and COVID-19 patients. *D-dimer* was increased in patients in specialized wards compared with patients in regular wards. *P-fibrinogen* was increased in patients in both regular and specialized wards compared with controls, but there were no differences observed between care levels. The data are also visualized in *Supplemental Figure S1* and *S2*.

**TABLE 2.** Laboratory test results for COVID-19 patients and healthy controls.

|  | COVID-19 |  |  | Healthy controls | Healthy controls versus COVID-19 patients (p) | Regular ward versus Specialized wards (p) |
| --- | --- | --- | --- | --- | --- | --- |
|  | Regular ward | Specialized wards | Total |  |  |  |
|  | N=40 | N=20 | N=60 | N=89 |  |  |
| Covid-positive, verified | 40 (100%) | 20 (100%) | 60 (100%) |  |  |  |
| D-dimer, mg/L, median (IQR) | 0.6 (0.5-1.0) | 1.5 (0.7-4.0) | 0.7 (0.5-1.5) |  |  | 0.002 |
| Hemoglobin, g/L, median (IQR) | 137 (125-148) | 123 (110-128) | 129 (120-141) | 141 (134-150) | <0.001 | <0.001 |
| Platelet count, 10<sup>9</sup>/L, median (IQR) | 212 (175-259) | 252 (206-341) | 221 (181-287) | 257 (229-283) | 0.005 | 0.094 |
| APTT, sec, median (IQR) | 26 (24-27) | 26 (25-30) | 26 (24-29) | 33 (31-36) | <0.001 | 0.891 |
| INR, median (IQR) | 1.0 (1.0-1.1) | 1.0 (1.0-1.1) | 1 (1-1.1) | 1.0 (1.0-1.1) | 0.107 | 0.843 |
| P-fibrinogen, g/L, median (IQR) | 5.4 (4.3-6.5) | 6.8 (4.8-7.6) | 5.7 (4.3-6.9) | 2.7 (2.4-3.0) | <0.001 | 0.125 |
| Antithrombin, kIE/L, median (IQR) | 1.0 (0.9-1.1) | 1.0 (0.9-1.1) | 1.0 (0.9-1.1) |  |  | 0.836 |
| EXTEM-CT, sec, median (IQR) | 70 (61-75) | 90 (78-108) | 74 (64-90) | 47 (43-51) | <0.001 | <0.001 |
| EXTEM-MCF, mm, median (IQR) | 70 (66-73) | 76 (71-77) | 71 (68-75) | 64 (62-68) | <0.001 | 0.009 |
| EXTEM-CFT, sec, median (IQR) | 49 (44-63) | 46 (42-55) | 49 (43-63) | 91 (79-101) | <0.001 | 0.485 |
| FIBTEM-MCF, mm, median (IQR) | 27 (23-31) | 32 (28-36) | 29 (24-33) | 13 (11-15) | <0.001 | 0.036 |

IQR=Interquartile Range. APTT=Activated Partial Thromboplastin Time. INR=International Normalized Ratio.

For EXTEM-CT, the Kruskal-Wallis test showed a significant difference between the groups (H(2) = 97.1, p < 0.001). Post-hoc pairwise Wilcoxon tests showed that COVID-19 patients (both care levels) had significantly longer CT compared with healthy controls (p<0.001) and that subjects treated at specialized wards had longer CT compared with subjects treated at regular wards (p<0.001) (*Figure 1A, Table 2*).

**Figure 1.**
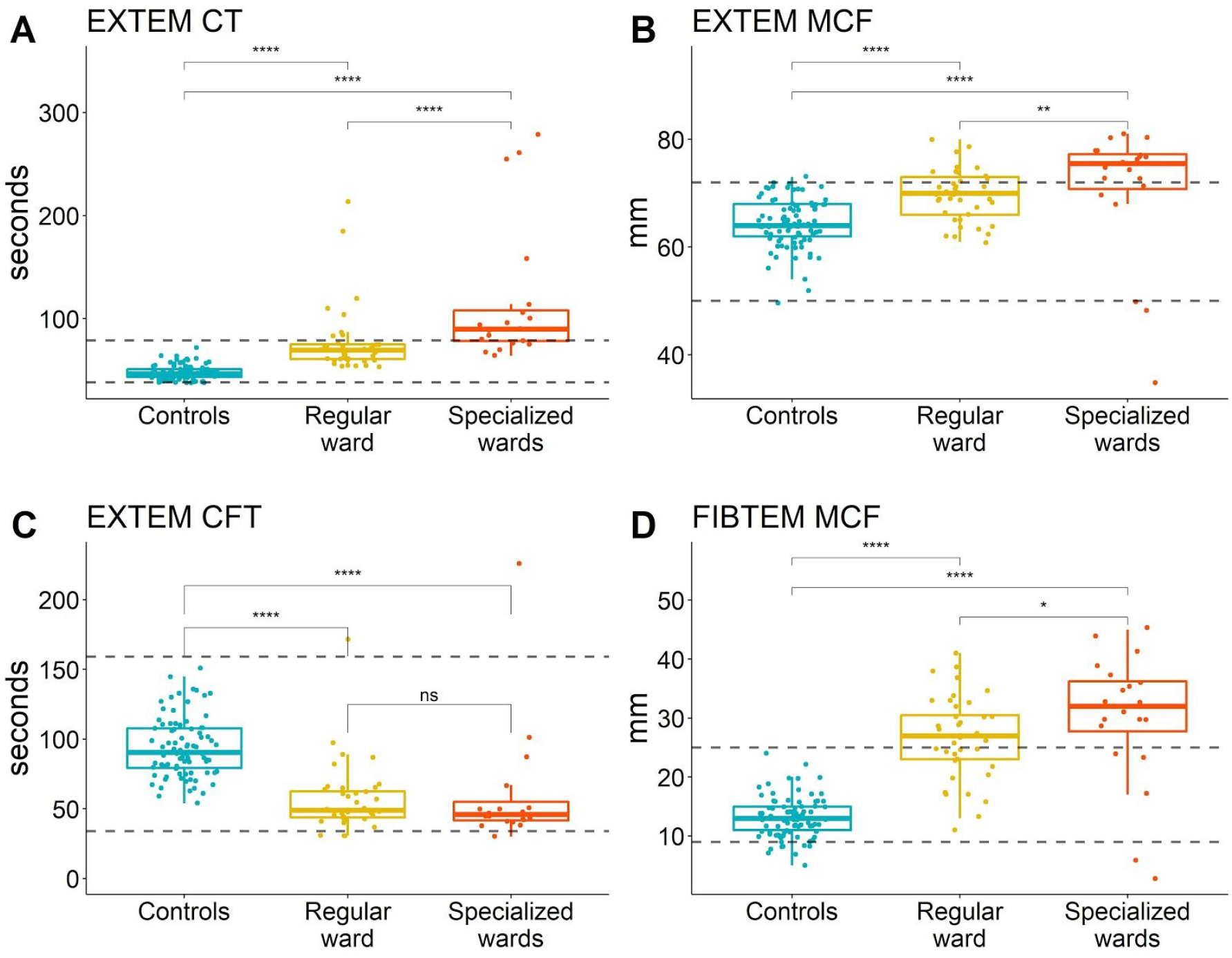
ROTEM data. Dashed horizontal lines are upper and lower reference values. p-values were calculated using a two-sided Wilcoxon signed rank test (ns = p > 0.05, ∗ = p < 0.05, ∗∗ = p < 0.01, ∗∗∗ = p < 0.001", ∗∗∗∗ = p < 0.0001). In A) EXTEM Coagulation Time (reference: 38-79 sec); B) EXTEM Maximum Clot Firmness (reference: 50-72 mm); C) EXTEM Clot Formation Time (reference: 34-159 sec); and D) FIBTEM Maximum Clot Firmness (reference: 9-25 mm).

For EXTEM-MCF, the Kruskal-Wallis test showed a significant difference between the groups (H(2) = 39.3, p < 0.001). Post-hoc pairwise Wilcoxon tests showed that COVID-19 patients (both care levels) had significantly increased MCF compared to healthy controls (p<0.001) and that subjects treated at specialized wards had increased MCF compared with subjects treated at regular wards (p<0.01) (*Figure 1B, Table 2*).

For EXTEM-CFT, the Kruskal-Wallis test showed a significant difference between the groups (H(2) = 64.8, p < 0.001). Post-hoc pairwise Wilcoxon tests showed that COVID-19 patients (both care levels) had significantly shorter CFT compared with healthy controls (p<0.001) (*Figure 1C, Table 2*).

For FIBTEM-MCF, the Kruskal-Wallis test showed a significant difference between the groups (H(2) = 79.5, p < 0.001). Post-hoc pairwise Wilcoxon tests showed that COVID-19 patients (both care levels) had significantly increased MCF compared with healthy controls (p<0.001) and that subjects treated at specialized wards had increased MCF compared with subjects treated at regular wards (p=0.04) (*Figure 1D, Table 2*).

To assess the association between D-dimer and EXTEM-MCF, and P-Fibrinogen and FIBTEM-MCF at an early stage of the disease, Spearman’s correlation coefficient was calculated for the subset of data where ROTEM data were available from the first day after hospital admission (N=34). There was no significant correlation observed between D-dimer and EXTEM-MCF (correlation = 0.02, p = 0.9). The correlation between P-Fibrinogen and FIBTEM-MCF was significant (correlation = 0.84, p < 0.001) (*Figure 2*).

**Figure 2.**
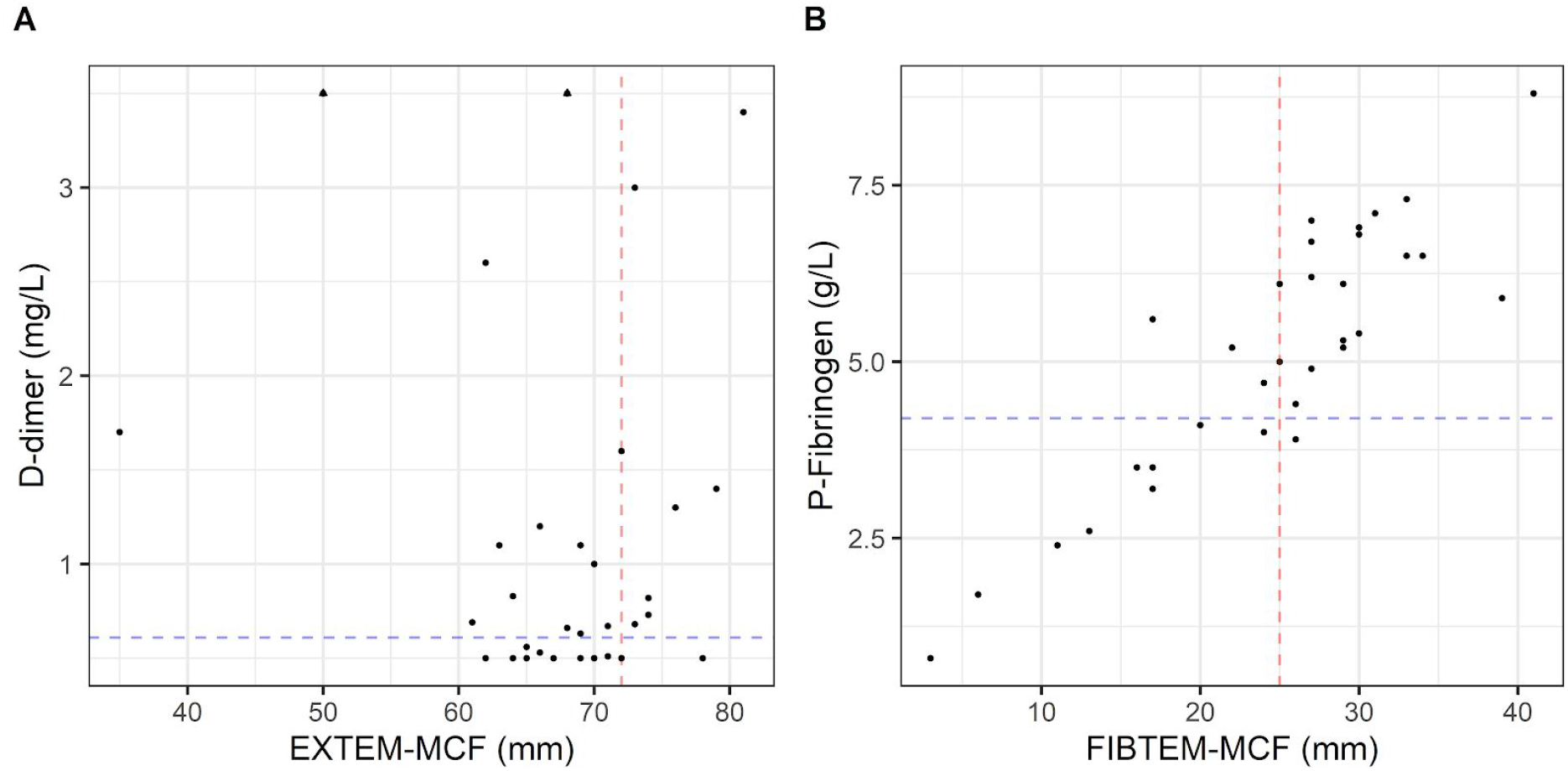
Scatter plots showing ROTEM data and coagulation markers for patients tested during the first day after admission. In A) D-dimer plotted against EXTEM-MCF. The red vertical line is the upper reference limit for EXTEM-MCF (72 mm); blue horizontal line is the upper reference limit for D-dimer (0.61 mg/L at age 61). Two D-dimer values higher than 3.5 are shown as triangles. In B) P-fibrinogen plotted against FIBTEM-MCF. The red vertical line is the upper reference limit for FIBTEM-MCF (25mm); blue horizontal line upper reference limit for P-Fibrinogen (4.2 g/L).

Our results show that the median patient with COVID-19 related mild to severe acute respiratory distress treated at StG between 200508-200531, had laboratory results indicating coagulopathy, with significant differences in ROTEM variables compared with healthy controls; longer EXTEM-CT, shorter EXTEM-CFT and increased EXTEM-MCF and FIBTEM-MCF. This suggests a patient with prolonged haemostatic initiation, shortened clot propagation and, notably, with a pronounced clot firmness, indicating hypercoagulation.

## Discussion

In this prospective observational study we assessed the level of coagulopathy in COVID-19 patients using ROTEM analysis. Previous studies have shown deranged values in critically ill patients ^9,10^. To our knowledge, this is the first time data on less severely ill patients early after admission to hospital are published. In our sample, we observed significantly higher EXTEM-MCF and FIBTEM-MCF in patients treated at regular wards compared with healthy controls. This indicates that coagulopathy is present early in the disease course and suggests that ROTEM-analysis may potentially be a useful predictor of thromboembolic complications and mortality.

During the months since COVID-19 was classified as a pandemic, evidence has emerged indicating coagulopathy and thrombosis as crucial explanations to why some patients develop severe illness ^2,3^. The pattern of higher EXTEM-MCF and FIBTEM-MCF in more severely ill patients in our data strengthens the hypothesis that the atypical respiratory symptoms observed in COVID-19 patients may partly be caused by thromboembolism impairing lung perfusion.

Increased D-dimer and P-fibrinogen have been reported to predict poor clinical outcome in COVID-19 patients ^6,7^. However, both these tests have caveats: the D-dimer blood test is nonspecific and may be increased in a variety of conditions including malignancy, inflammation and infection ^13^; fibrinogen is an acute phase reactant, and high levels of P-fibrinogen may reflect a patient with a high inflammatory profile, which itself may amplify the effects of other cardiovascular risk factors ^14^. Furthermore, conventional coagulation tests (CCTs) are limited by their inability to assess clot strength, fibrinogen functionality and fibrinolysis ^15^. Conversely, ROTEM provides a more rapid and comprehensive assessment of whole blood clot formation allowing for a complete view of the entire coagulation cascade, and has been shown a better tool for monitoring coagulation profiles than CCTs ^16,17^.

As can be seen in *Figure 2*, FIBTEM-MCF and P-fibrinogen are highly correlated in COVID-19 patients, whereas EXTEM-MCF supplies information that differs from D-dimer at an early disease stage. If individuals at risk of developing COVID-19 related thromboses are identified at an early stage, enhanced prophylaxis with LMWH may decrease mortality in this group of patients ^6 18^. It is feasible that ROTEM could be applied for this purpose. Whether the ROTEM data will have better predictive characteristics compared with CCTs do, however, remain to be shown.

Sepsis is known to exert a complex effect on the coagulation system, generally affecting all aspects of clot formation, kinetic activity and clot development ^19^. Already in the early stages of sepsis the coagulation system and the platelets are strongly activated ^20^. However, it has been recently shown that the coagulopathy related to the COVID-19 pathogenesis differs from the disseminated intravascular coagulation (DIC) associated with sepsis, with relatively normal levels of INR, APTT and platelets despite markedly elevated d-dimer levels ^21 22^. Our results confirm these findings. Nevertheless, whether this hypercoagulability is due to the characteristics of the invading microorganism, the individual viral load, or the massive host inflammatory response remains unknown. The coagulopathy related to COVID-19 respiratory distress may in some way be specific for the SARS-CoV-2 species, representing new features that need to be further explored.

## Limitations

Our study has limitations. First, 60% of patients had received anticoagulant treatment before ROTEM analysis, usually low dose, which may have to some degree counteracted hypercoagulation. However, despite this limitation, ROTEM results were highly significantly increased in COVID-19 patients compared with healthy controls. Second, not all patients admitted for COVID-19 at St Göran’s Hospital (StG) during this period were tested using ROTEM, and not all patients that were included were tested during the first day after admission. The main reasons for this were: (i) Due to clinical duties the physicians responsible for the study were not always able to scan the wards for new patients on a daily basis (ii) and more importantly, limitation of the testing equipment, with possibility to run a maximum of 6 tests every 4 hours. Still, we believe that the included patients are a representative cross-section of the COVID-19 patients treated at StG during the study period.

## Conclusion

In hospitalized COVID-19 positive patients, elevated values of EXTEM-MCF and FIBTEM-MCF were seen at admission to hospital, suggesting a hypercoagulable state. This pattern was more pronounced in patients with more severe disease. Whether ROTEM-analysis at admission to hospital has value as a predictor of clinical outcome (e.g., mortality, thrombosis) in COVID-19 positive patients will be analyzed at a later stage of this research project.

## Data Availability

Data and code will be made available upon reasonable request

## Acknowledgements

We wish to thank the staff at the COVID-19 wards and the Laboratory Unit (especially Jacqueline Akcan and Charlotta Törnberg) at St Göran’s Hospital for collaboration and involvement in this study, and colleagues at the Intensive Care Unit for support. Special thanks to Rasmus Berglind, Anton Borgström and Christine Carlswärd for valuable assistance with data collection.

## Authors’ contributions

All authors have made substantial contributions to this study. LA, AW, JS, MW, MB, JvdL and AÅ conceived and designed the study. LA made the data collection. JS performed the data analyses and visualizations, and MB contributed to the data analysis. LA and JS drafted the manuscript. AW, MW, JvdL and AÅ provided input for interpretation of results and clinical expertise. All authors have read, critically revised and approved the final manuscript.

## Conflict of Interests

The authors declare no conflict of interests.

## Funding

This work was supported by an unrestricted grant from CSL Behring (AW) and grants from Karolinska Institutet (JvdL).

## Notes

### Competing Interest Statement

The authors have declared no competing interest.

### Funding Statement

This work was supported by an unrestricted grant from CSL Behring (AW) and grants from Karolinska Institutet (JvdL).No other author report external funding for the study.

### Author Declarations

The study was approved by the Swedish Ethical Review Authority (D-nr 2020-01875).

